# Same-Day Simultaneous Diagnosis of Bacterial and Fungal Infections in Clinical Practice by Nanopore Targeted Sequencing

**DOI:** 10.1101/2020.04.08.20057604

**Authors:** Ming Wang, Aisi Fu, Ben Hu, Gaigai Shen, Ran Liu, Wanxu Zhao, Shupeng Jiang, Xuan Cai, Congrong Li, Juan Li, Qing Wu, Kai Feng, Jiashuang Gu, Jia Chen, Mingyue Shu, Binghong Zhang, Zixin Deng, Lilei Yu, Yan Li, Tiangang Liu

## Abstract

**BACKGROUND:** As approximately 19% of global deaths are attributable to infectious diseases, early diagnosis of infection is very important to reduce mortality. Traditional infection detection strategies have limited sensitivity, detection range, and turnaround times; a detection technology that can simultaneously detect bacterial and fungal infections within 24 h is urgently need in clinical settings.

**METHODS:** We developed nanopore targeted sequencing (NTS) for same-day simultaneous Diagnosis of fungal and bacterial infections. NTS was developed by amplification of 16s rRNA gene (for bacteria), IST1/2 gene (for fungal), and *rpoB* (for *Mycobacterium* spp.) using multiple primers, and sequenced by a real-time nanopore sequencing platform. An in-house bioinformatic analyze pipeline was used to diagnose the infectious pathogens by mapping the sequencing results with the constructed databases.

**RESULTS:** Comparison of 1312 specimens from 1257 patients using NTS and culture method; NTS detected pathogens in 58.71% of specimens from patients, compared to 22.09% detected using the culture method. NTS showed significantly higher sensitivity than culture methods for many pathogens. Importantly, a turnaround time of <24 h for all specimens, and a pre-report within 6 h in emergency cases was possible in clinical practice. Modification of antibiotic therapy and maintenance of original anti-infection regimens in 51.52% (17/33) and 36.36% (12/33) of patients was in accordance with NTS results, and quantitative monitoring of clinical treatment effects was evaluated in four patients by continuous NTS tests.

**CONCLUSIONS:** Application of NTS in clinically detected pathogens can improve targeted antibiotic treatment and therapeutic monitoring.

## Introduction

Approximately 19% of global deaths are attributable to infectious diseases, and early empirical treatment may not improve survival rates or other outcomes *(1)*. The low sensitivity or biased detection range of individual techniques results in the combined use of pathogen screening techniques, which will lead to repeated sampling, increased treatment costs, and extended diagnosis time *(2-4)*. Furthermore, the need for rapid pathogen identification, a wide pathogen spectrum *(3)*, limited specimen volumes *(5)*, or even asymptomatic infections *(6)* should be considered.

Enormous progress has been made in the development and deployment of sequencing-based diagnostic methods *(4, 7-10)*. Sequencing-based diagnostic methods have broad detection ranges and high resolution, particularly for fungal pathogens. However, unbiased metagenomic methods typically require deep sequencing and extensive bioinformatics processing to account for host genome dominance, resulting in low-throughput, high costs, long turnaround times, and low sensitivity *(4, 11)*. Targeted sequence detection requires less sequencing data and bioinformatics resource, which in theory, can reduce the cost and turnaround time, therefore suitable for clinical applications. However, due to the short-read lengths of previous sequencing platforms, its discrimination of pathogens is limited by incompletely sequenced marker genes, such as the V4-5 region of 16s rRNA gene. The detection range was also restricted to only bacteria or fungi in a single test due to their different sized marker genes which cannot be constructed in same sequencing library *(8, 12)*.

DNA is sequenced by massively parallel sequencing platforms by detecting optical or chemical signals. This method accumulates systematic errors generated by sequencing as the sequencing length increases, limiting the length of sequencing reads *(13, 14)*. Illumina’s (the most widely used massive parallel sequencing platform) sequencing by synthesis approach requires multiple sequencing cycles, and each cycle takes several minutes to analyze one base of each DNA fragment. Hence, the sequencing process usually takes 0.5 to 3 days according to the read length and data output requirement. More importantly, the sequencing data cannot be used for following analysis until the entire sequencing process is completed. Nanopore sequencing platform’s approach directly detects changes in the generated current when DNA / RNA molecules pass through a nanopore protein at approximately 300 bp/s, which now achieving >95% raw read accuracy *(15)*. This electrical signal can be recorded in real-time and used for subsequent sequence analysis immediately *(16)*. In addition, the nanopore protein does not selectively sequence DNA / RNA with specific lengths. DNA / RNA of different lengths can pass through the nanopore protein completely, and the length of each sequencing data is equal to that of the length of the DNA / RNA *(17)*. Therefore, unlike previous sequencing platforms, the length of nucleic acids in the same sequencing library is not required to be of a specific range, such as 280-320 bp; all nucleic acids with different length can be pooled together and sequenced from end to end in a nanopore sequencing library *(18, 19)*.

Real-time data output, end to end sequencing and small size make nanopore sequencing promising for clinical applications. Previous studies focused on bacterial respiratory infection using nanopore metagenome sequencing *(20-22)*, however the clinical application was impeded by relative high costs and extensive bioinformatics processing. Nanopore targeted sequencing (NTS), which amplified marker genes and used nanopore sequencing platform to sequence the amplified marker genes, were developed for investigation of bacterial or fungal infection. However, previous NTS was only based on a single marker gene, such as the 16s rRNA gene targeted sequencing that has been reported to identify bacterial infection in individual clinical case reports *(23, 24)*, and fungal ribosomal operon (18S-ITS1-5.8S-ITS2-28S) has been used to detect the fungal community associated with external otitis in dogs *(25)*. Simultaneous broad-spectrum detections of bacteria and fungi, and high-sensitivity detection of critical clinical pathogens in clinical samples by a single test based on NTS have not been developed. In addition, the performance of the NTS method with a large cohort and systematic comparisons with classic culture methods have not been examined. Furthermore, the turnaround time in clinical practice and clinical management applications are unknown.

Here, we developed a test based on NTS for the broad-spectrum screening of both bacterial and fungal pathogens in various clinical specimen types. The diagnostic accuracy and turnaround time of NTS were investigated by comparison with the culture method for 1312 clinical specimens from 1257 patients.

## Materials and Methods

### SPECIMENS AND CLINICAL ELECTRONIC RECORD

The study population consisted of 1312 specimens from 1257 patients at Renmin Hospital of Wuhan University, China, whose specimens were sent for pathogen testing between November 2018 and November 2019. Parallel testing by routine culture and NTS was performed. Adequate material was submitted for culture, and surplus material was processed by NTS. All specimens (n = 1312) were used to assess the NTS and culture results. The clinical records of each patient were retrieved and 1005 patients were diagnosed or suspected with infection by clinicians according to clinical symptoms and/or positive results of other clinical tests. These patients were used to evaluate the diagnostic yield and sensitivity of NTS. Patient clinical diagnosis and demographic characteristics were obtained from electronic medical records *(26)*. The study was approved by the Ethics Committee of Renmin Hospital of Wuhan University (WDRY2019-K056).

### NTS

All primers used in this study are listed in Table S1, the DNA extraction method of specimen was described in supporting method. Amplification of the 16s rRNA gene and *rpoB* was performed in a 20 μL reaction system with 8 μL extracted DNA, 2 μL barcoded primer (10 μM), and 10 μL 2×KOD OneTM PCR Master Mix (TOYOBO) using the following procedure: 1 cycle at 98°C for 3 min and 35 cycles at 98°C for 10 s, 55°C for 5 s, and 68°C for 10 s, followed by a final elongation step at 68°C for 5 min. ITS1/2 (fungal internal transcribed spacers 1 and 2, abbreviated as ITS1/2) was first amplified using the same reaction system and PCR procedure using the primer mix without barcode and the PCR product was purified with 0.8× AMpure beads (Beckman Coulter) and eluted in 10 μL Tris-EDTA (TE) buffer. Then, 5 μL eluate was used for barcoding PCR with 5 μL barcoded ITS1/2 primer set (10 μM) and 10 μL 2× Phusion U Multiplex PCR Master Mix using the following procedure: 1 cycle at 98°C for 3 min and 10 cycles at 98°C for 10 s, 55°C for 5 s, and 68°C for 5 s, followed by a final elongation step at 68°C for 5 min. The barcoded products of the 16s rRNA gene, ITS1/2 and *rpoB* amplification from the same samples were pooled according to mass ratio of 10:3:1. The pooled products from the different samples were mixed equally and used to construct sequencing libraries using the 1D Ligation Kit (SQK-LSK109; Oxford Nanopore). The library was sequenced using Oxford Nanopore MinION or GridION. TE buffer was assayed in each batch as a negative control. The bioinformatic analysis was described in the supplement methods section, several modifications were made following previous studies and samples were tested positive for bacteria or fungi if they met any of the established thresholds (Supporting method). Detected microorganisms were described as critical pathogens, opportunistic pathogens, or typically nonpathogenic commensal microbes according to published literature and clinical guidelines *(27)* (Table S2).

### MOCK COMMUNITY

All 50 clinical strains used to evaluate performance were isolated from clinical specimens in the laboratory or purchased from the American Type Culture Collection (ATCC) (Manassas, VA, USA). Six strains were selected to construct a mock community (Table S3) and blood samples were collected from healthy volunteers with no symptoms of infection in physical examination. The concentration of each strain was determined by calibration curves from cultures.

### STATISTICAL ANALYSIS

The sensitivity, specificity, positive predictive value (PPV), and negative predictive value (NPV) were calculated as previously described by comparing the NTS results with composite final clinical diagnosis *(28)*. A P-value < 0.05 was considered significant. Proportional outcomes were compared using the ^2^ test or Fisher’s exact test. Continuous variables were compared using the Student’s *t*-test or Wilcoxon signed-rank test. Data analyses were performed using SPSS 22.0.

## Results

### DESIGN OF NTS FOR SAME-DAY SIMULTANEOUS DIAGNOSIS OF BACTERIAL AND FUNGAL INFECTIONS

Full-length bacterial 16s rRNA gene and fungal ITS1/2 gene were selected as universal marker genes for identification of bacteria and fungi, respectively. For specific markers, as *Mycobacterium tuberculosis* (MTB) was the most concerned clinical critical pathogen, we chose *rpoB* for *Mycobacterium* spp. *(29)* as the target. The design of NTS primers (Table S1) and the sequencing library preparation were described in the supporting result. For sequencing, we chose a nanopore platform that could sequence long nucleic acid fragments and simultaneously analyze the data-output in real-time. This allowed immediate detection of bacterial and fungal infection by periodical analysis of output sequences using our in-house bioinformatic pipeline (supporting methods). In summary, a NTS detection that targeted the amplification of bacterial and fungal marker genes and that is able to execute nanopore sequencing to identify the pathogens was developed (Fig. 1).

**Fig. 1.**
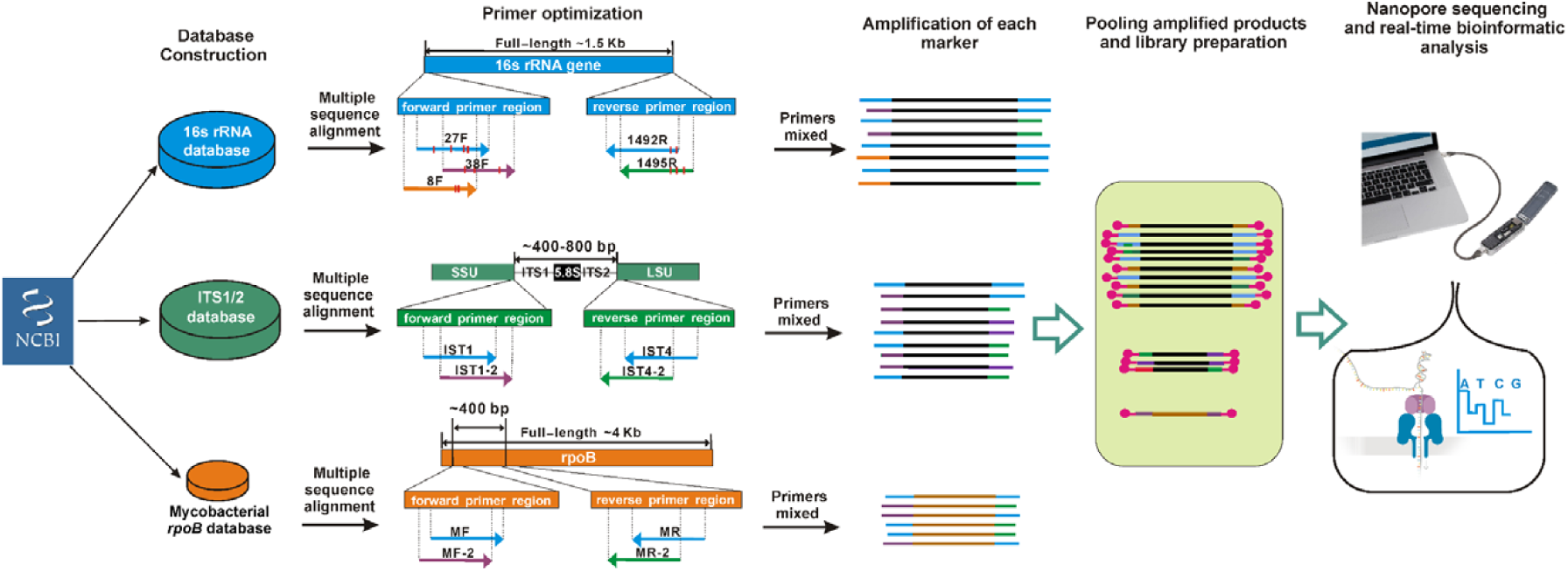
Design of NTS for simultaneous detection of bacterial and fungal infection. Three marker gene databases, corresponding to 16s rRNA, ITS1/2, and *rpoB*, were constructed from the sequences from the NCBI database. The results of multiplex sequence alignment of each database was used to analyze the conservative region and degeneration of each marker gene base. The blue primers were used in previous studies and selected as “start primers” for the design of other “additional primers” which is labeled by other colors. The red vertical bar on the primer indicates degenerate base. The “start primers” and “additional primers” for each marker gene was mixed at the molar ratio of 3:1. The mixed primers were then used to amplify the marker gene, individually. The PCR products of the 16s rRNA, ITS1/2, and *rpoB* from the same sample were then pooled at a mass ratio of 10:3:1. This pooled product was used for library preparation, nanopore sequencing and bioinformatic analysis.

### PERFORMANCE OF NTS BASED ON STANDARD STRAINS AND MOCK COMMUNITIES

To verify the identification accuracy, NTS was tested by individual analyses of 50 common clinical strains isolated from clinical specimens in the laboratory or purchased from ATCC. In total, 100% and 92% of the tested isolates were identified correctly at the genus and species level, respectively (Table S4). Considering clinical samples, such as sputum, that usually contain high abundance of commensal microbes and that the pathogens may have relatively lower abundance, in order to investigate the sensitivity and sequencing data required for detection the low-abundance microbes, a mock community (three bacteria and three fungi) was constructed to simulate a complex clinical sample and the concentration of each strain was determined by calibration curves from cultures (Table S3). Considering that different levels of background human DNA could potentially affect sensitivity, the performance of NTS was tested using healthy human blood spiked with varied concentrations of the mock community. NTS detected each species at all six pathogen densities, and the least abundant fungi and bacteria, with 25 colony-forming units (CFU) in a community of 1000 CFU, were consistently detected in 12 repeats (Fig. 2A, B). Therefore, the limit of detection was determined to be 25 CFU in 12/12 positive replicates (Table S5).

**Fig. 2.**
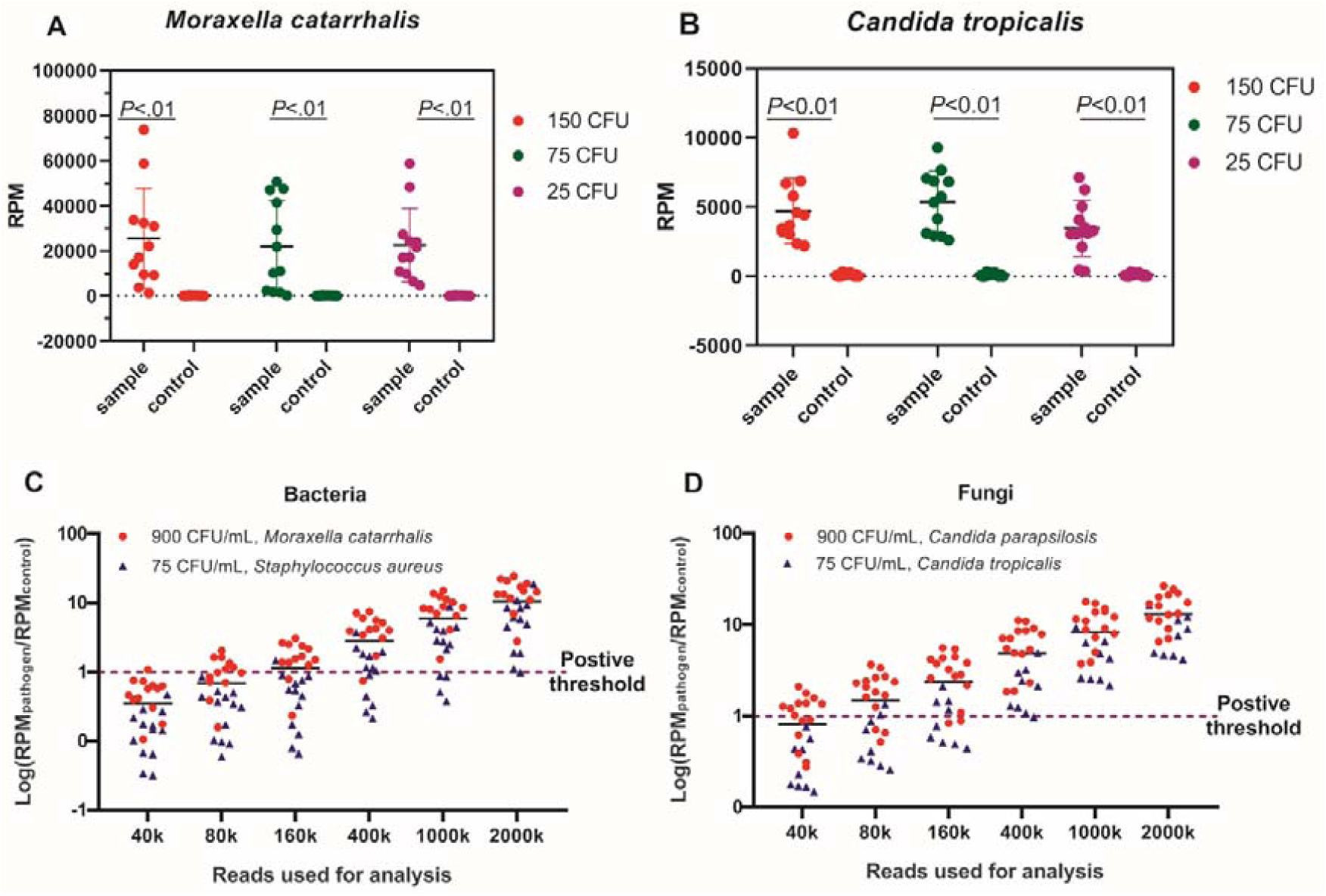
Proficiency test of NTS with a mock community. Detection of the least abundant bacteria (A) and fungi (B) in the mock community. *Moraxella catarrhalis* and *Candida tropicalis* each accounted for 5% of the mock community. Three different densities from 25 to 150 CFU/mL were tested with 12 repeats. Each spot represents a repeat detection. (C) Comparison of the RPM of each strain in the mock community in a saline background with that in human blood. The correlation between the detection of bacteria (C) and fungi (D) with sequencing data used for the analysis. Dashed line indicates the positive threshold between the sample and control used as the limit of detection for pathogens in this study. Abbreviations: NTS, nanopore targeted sequencing; CFU, colony-forming units; RPM, aligned reads of per million sequence reads.

To determine a proper sequencing time (corresponding to sequencing data) that is required for confident detection of different amounts of microbes in community, a sequencing library contained those 48 samples was sequenced on a chip for 12 h. The most abundant bacteria or fungi, accounting for 30% of the mock community, were detected in all 12 repeats using total 400,000 sequencing reads, which required approximately 30 min. Meanwhile the least abundant bacteria or fungi, accounting for 5% of the mock community, were identified using total 2,000,000 reads, which required approximately 4 h (Fig. 2C, D). This indicated that 4 h sequencing was enough for detection of all pathogens and that a sequencing library with fewer samples could reduce the sequencing time. Considering the diversity of clinical samples, in which pathogens may account for >30% or <5% of the microbial community, the pathogen detection time can be achieved <30-minute or require > 4 h of sequencing time.

### COMPARISON OF NTS AND CULTURE METHOD IN CLINICAL PRACTICE

Upon verification of the performance of NTS using the mock community, 1312 clinical specimens from 1257 patients were parallelly analyzed by NTS and culture, including 301 and 1011 specimens from sterile (low commensal microbes background) and non-sterile settings (high commensal microbes background), respectively.

Using the culture-based results as a reference, among 289 culture-positive cases, NTS detected pathogens in 97.58% cases (n = 282/289), including identical findings to culture results in 52.94% cases (n = 153/289), detection of additional potential pathogens other than the culture results in 30.44% of cases (n = 88/289), different species of same genus detected between NTS and culture (6.92%, n = 20/289) and different pathogens detected between two methods (7.27%, n = 21/289) (Fig. 3A). Considering the different growth rates of bacteria and fungi, 84 of the 88 cases of additional pathogens detected by NTS were diagnosed as bacterial or fungal infections only by culture methods, in which additional critical pathogens (n = 2), additional opportunistic fungi or bacteria (n = 67), and potential coinfection with opportunistic fungi and bacteria (n = 19) were detected (Fig. 3A). The NPV of NTS (culture as reference) was 98.88%, which indicated that NTS was able to detect most of the pathogens identified by culture (Table 1). The pathogens detected by culture methods were missed by NTS in 2.42% cases (n = 7/289), in which high background of commensal microbes may impact the detection of pathogen with low abundance.

**Table 1.**
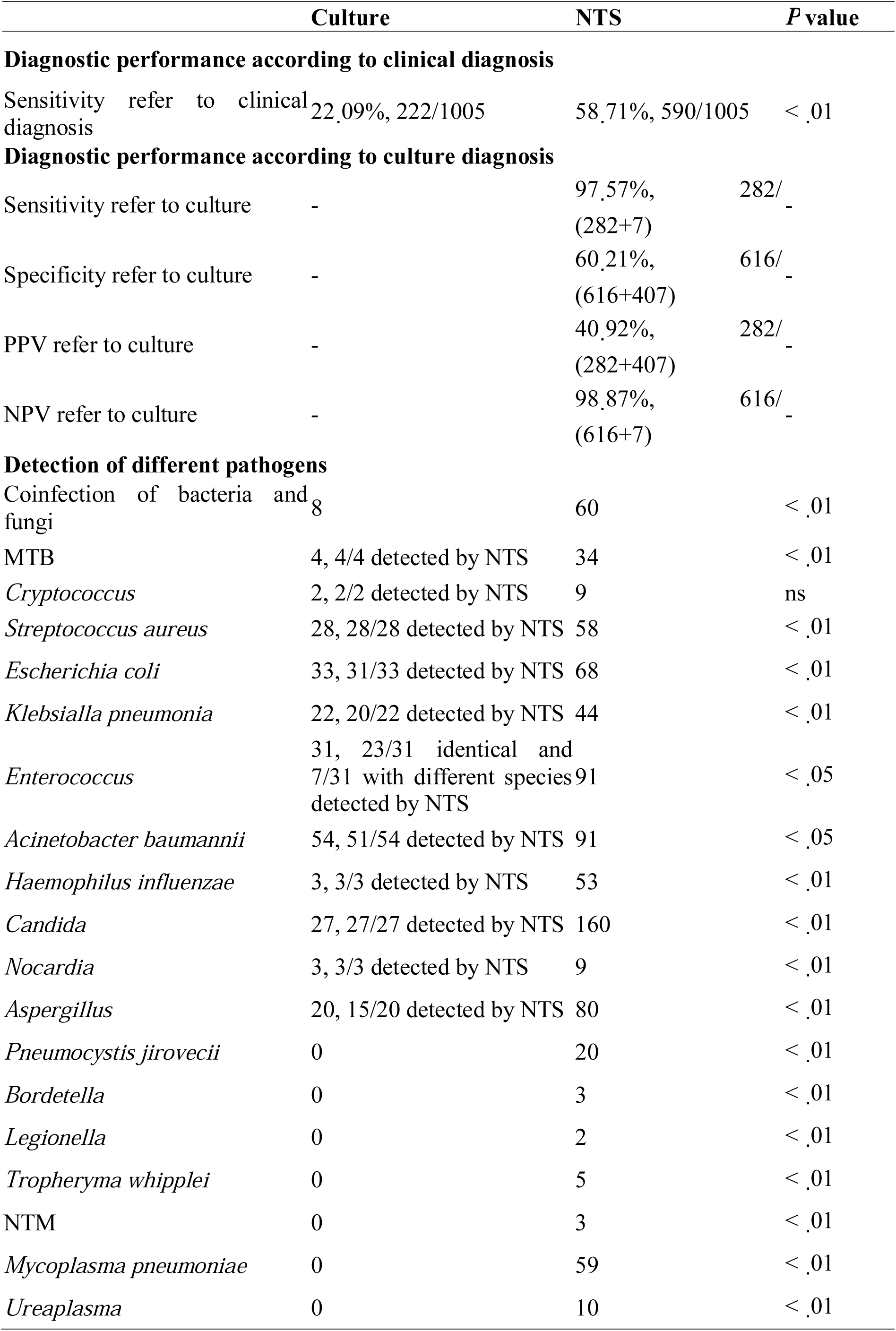
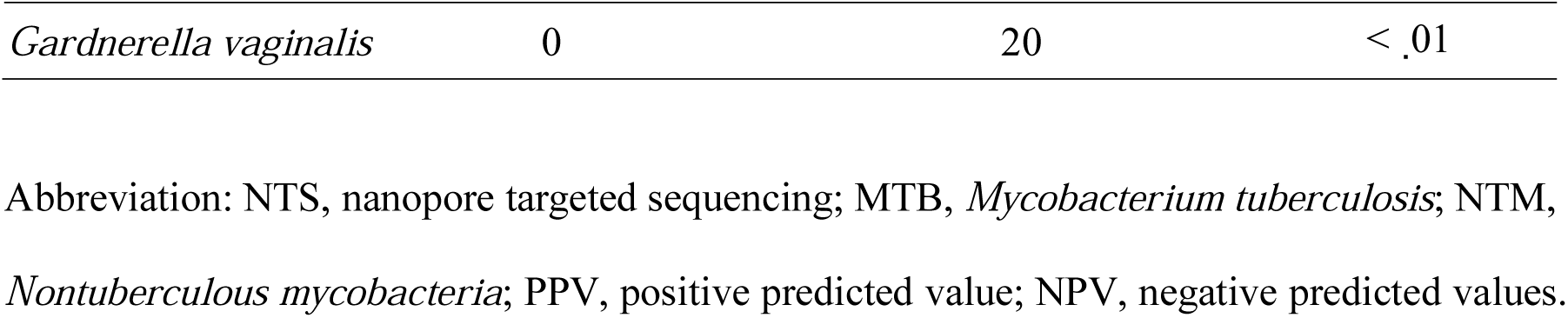
Diagnostic Performance of NTS and Culture.

**Fig. 3.**
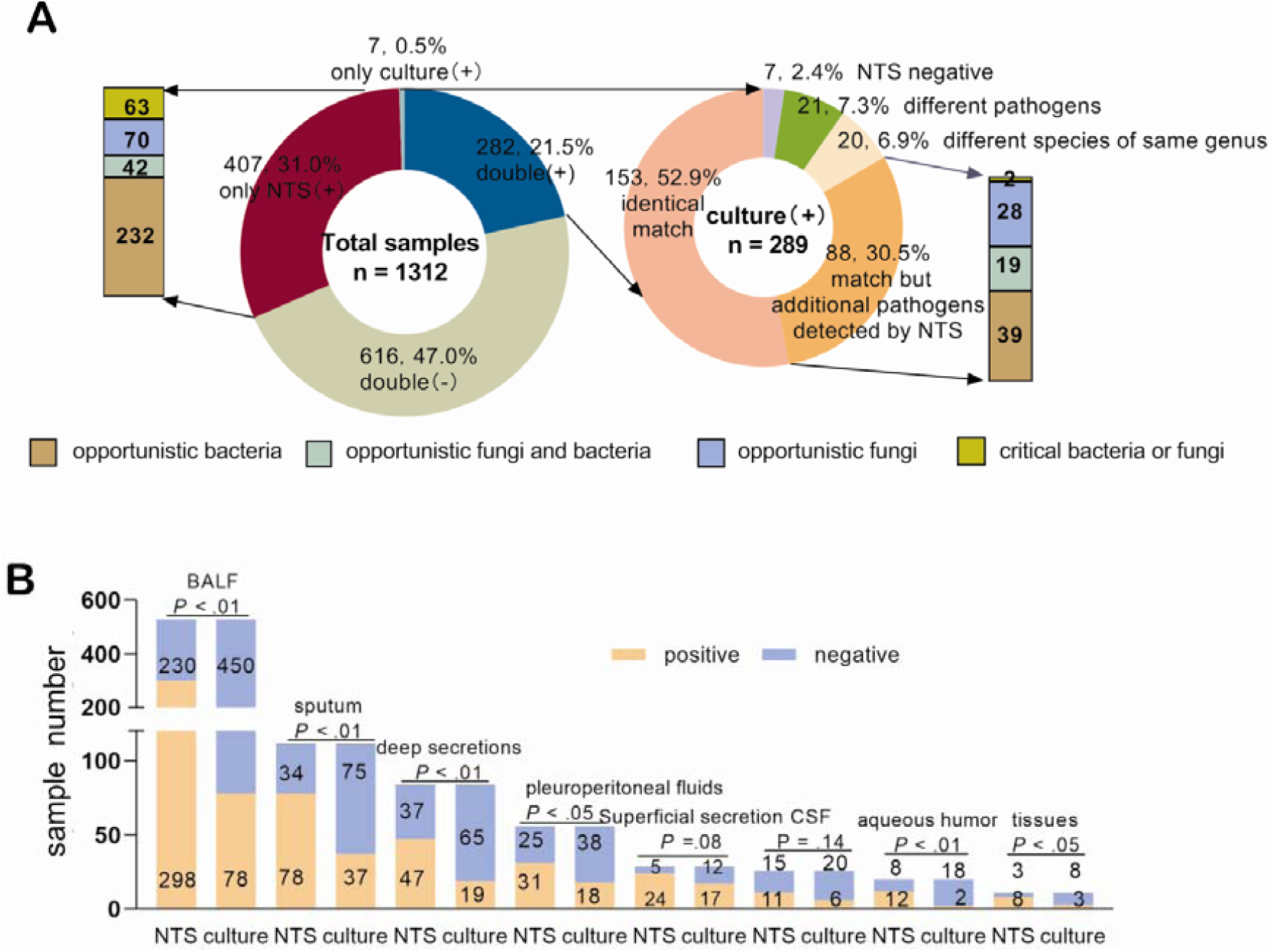
Comparison of NTS and culture method in real clinical practice. (A) Evaluation of concordance between NTS and culture results. Detected microbes were defined as critical or opportunistic bacteria or fungi according to previous studies and clinical guidelines. (B) Positive rates of NTS for each kind of specimen compared to that of culture results using the clinical diagnosis of each patient as reference. Abbreviations: NTS, nanopore targeted sequencing; BALF, bronchoalveolar lavage fluid; CSF, cerebrospinal fluid.

Of the 1023 specimens identified as culture-negative, potential pathogens were detected in 39.78% cases (n = 407/1023) by NTS, especially including 63 critical pathogens (Fig. 3A). In order to evaluate the accuracy of the NTS results of these cases, GeneXpert (a Food and Drug Administration-approved assay based on specific PCR of *rpoB* in MTB) analysis was conducted in 27/34 MTB-positive patients determined by NTS, and 100% concordance was observed; the other 7 patients were confirmed by acid-fast staining (n = 2) or transferred to a specialized hospital according to the clinical symptoms without further confirmation (n = 5) (Table S6).

According to the clinical diagnosis record around the time of the parallel NTS and culture test, 1005/1257 patients were diagnosed or suspected as infection by clinicians according to the clinical symptoms and/or positive results of other clinical tests. However, culture only reported 289 positive cases, which indicated that several pathogens could have been missed and could not to be a reference for comprehensive assess of NTS. Thus, clinical diagnosis was used to evaluate the performance of NTS in infection diagnosis and practicability in clinical application. The sensitivity of NTS (diagnose as reference) were 58.7% (n = 590/1005), which was >36% higher than that for culture-based methods (22.1%, n = 222/1005, *P* < 0.01), and 52 more potential coinfections with bacteria and fungi were detected by NTS (Table 1). For slow-growing, fastidious pathogens, NTS exhibited a significantly higher positive detection rate compared to that of culture methods, particularly for MTB (34 vs. 4), *Aspergillus* (80 vs. 20), *Pneumocystis jirovecii* (20 vs. 0), and *Nocardia* (9 vs. 3) (Table 1). Moreover, considering the antibiotic pre-exposure in nearly all patients from the Departments of Respiration and Neurology, no colonies, including fast-growing oral flora and colonizers, grew after a 72-h incubation period in 44/401 and 55/62 specimens, among which NTS showed 40.9% (n = 18/44) and 21.0% (n = 12/55) positive results. In those NTS-positive samples, 11/18 (61.1%) and 10/12 (83.3%) were culturable pathogens, such as *Streptococcus aureus* (Table S7).

Specimen volume, types of pathogens, abundance of colonizing microorganisms, and concentration of human DNA backgrounds in different clinical specimens existed to a large diversity. To assess the impact of different types of specimens on the efficiency of NTS testing, we evaluated the sensitivity of NTS in different specimens. The positive rate of NTS was higher than that of culture methods, particularly in the aqueous humor, which usually has limited specimen volume (60.0% vs. 10.0%, *P* < 0.05), and bronchoalveolar lavage fluid (BALF), which usually contains high abundance of commensal microbes (56.4% vs. 14.8%, *P* < 0.05), except for superficial secretions (*P =* 0.08) in which the main pathogens were fast-growing bacteria, such as *Streptococcus aureus* (Fig. 3B). These results indicated that detection sensitivity of NTS was less impacted by the type of specimen and that NTS was suitable for the infection detection of various specimens.

For all specimens, considering the complexity of clinical samples and more sequencing data may improve sensitivity, the default total sequencing time was defined as 8 h; the time from sample collection to the generation of the pathogen report was within 24 h for NTS (n = 1312), which was significantly shorter than the estimated median time of 72 h for culture methods (P < 0.001) (Fig. 4). Notably, real-time information was generated during sequencing, and a pre-report can be obtained if the data meet the positive criteria, further reducing the turnaround time. Briefly, in many emergency cases, 10 min sequencing data was sufficient to identify the pathogens and targeted antibiotics can be administrated by clinicians on the same day of detection without waiting for the completion of an 8 h sequencing.

**Fig. 4.**
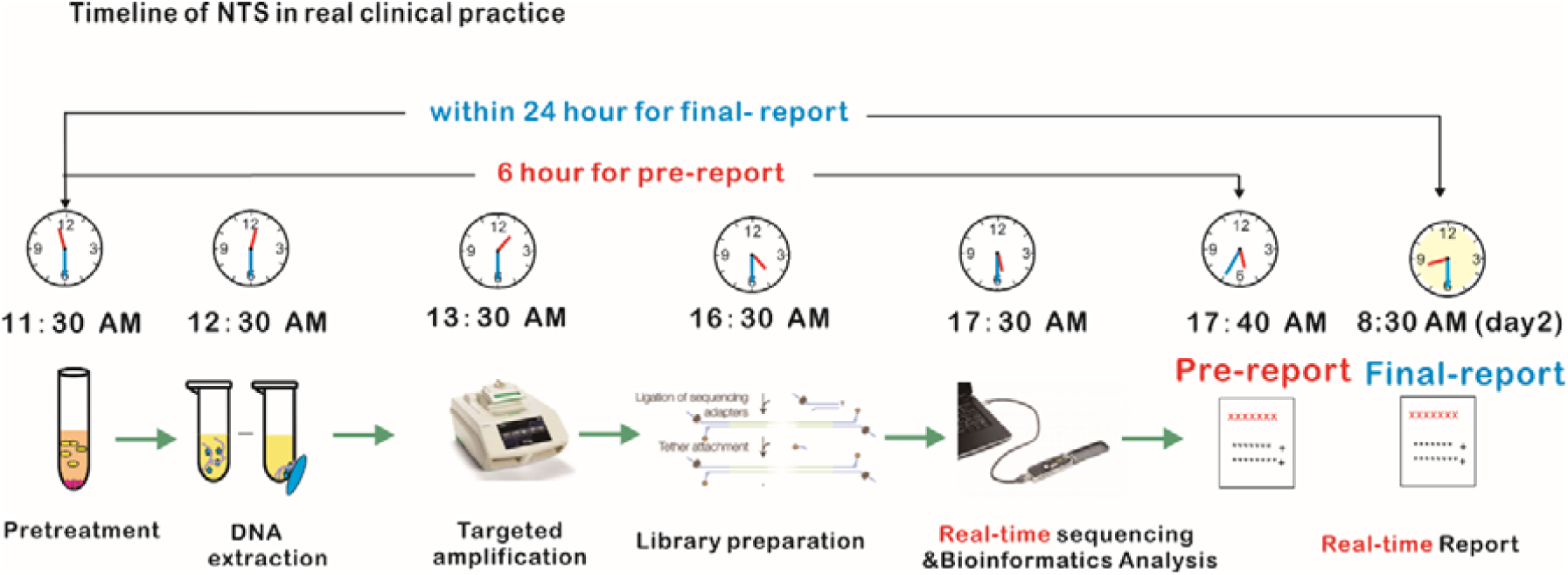
Schematic illustration of the total duration of NTS in clinical practice, including pretreatment, DNA extraction, targeted amplification and library construction, sequencing, bioinformatic analysis, and generation of the real-time report.

### EFFECT OF NTS ON ANTIBIOTIC MANAGEMENT

To assess the impact of pathogen detection by NTS on antibiotic management, we investigated the patients’ electronic records. Empirical antimicrobial agents were initiated before NTS in 93.9% (31/33) of NTS-positive samples in the Department of Respiratory ICU. Based on the NTS results, 51.52% (17/33) of patients were switched to targeted antibiotics (n = 7) or different antibiotics covering the same antibacterial spectrum (n = 10). NTS results did not trigger modification of antibiotic therapy in 42.42% (14/33) of patients; among these patients, 85.71% (12/14) maintained the original anti-infection regimen, which already covered the pathogen detected by NTS, and 14.29% (2/14) were discharged from hospital without subsequent therapy (Table S8). Four patients were evaluated by continuous NTS assay during the treatment process. In four cases, the pathogens detected in the first NTS assay and triggering antibiotic modification were eliminated according to the results of the second NTS assay, resulting in the suspension or de-escalation of targeted antibiotics. In one case, other pathogen were detected in the second NTS, which may have been transient colonizers with the potential to induce a nosocomial infection (Table 2).

**Table 2.**
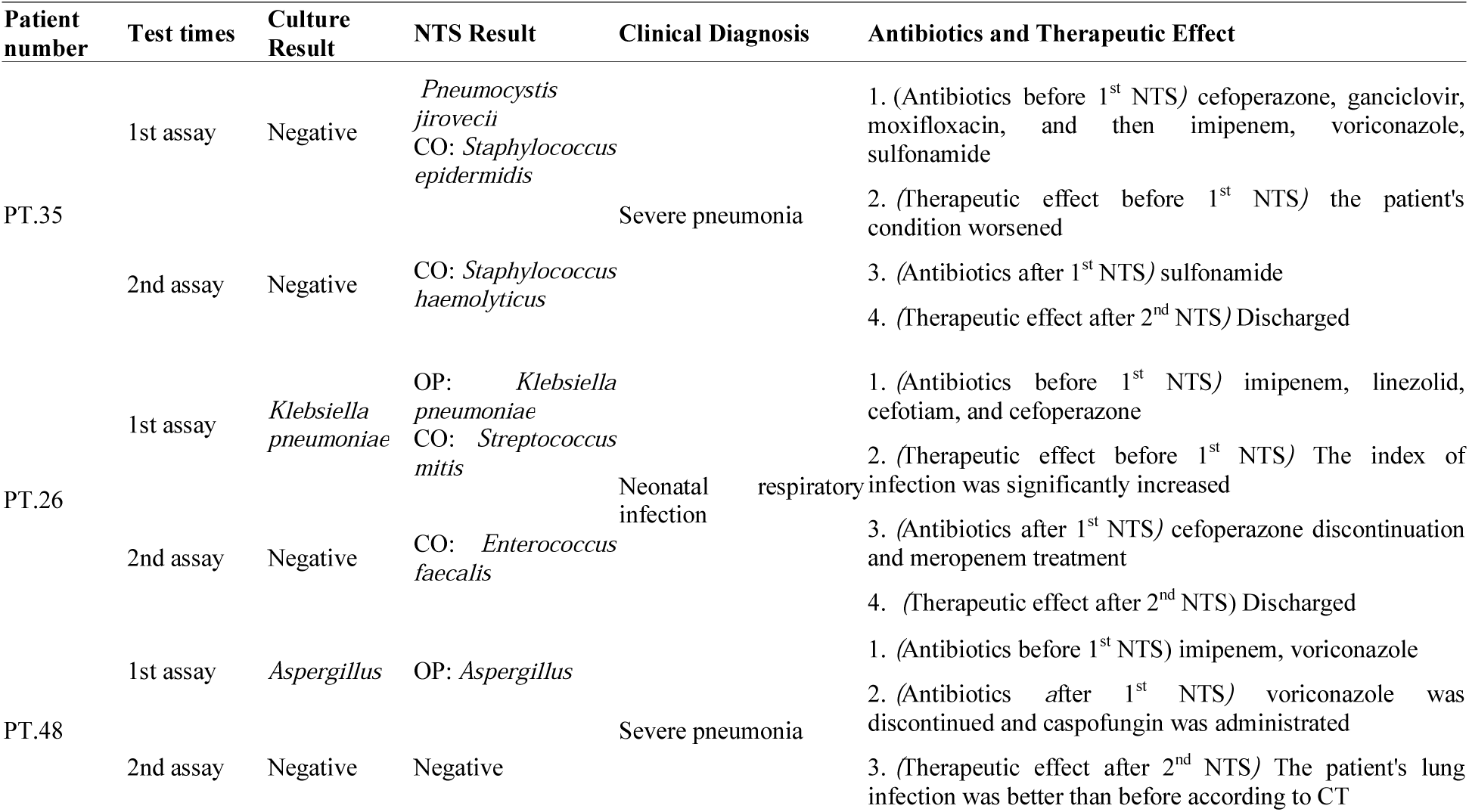
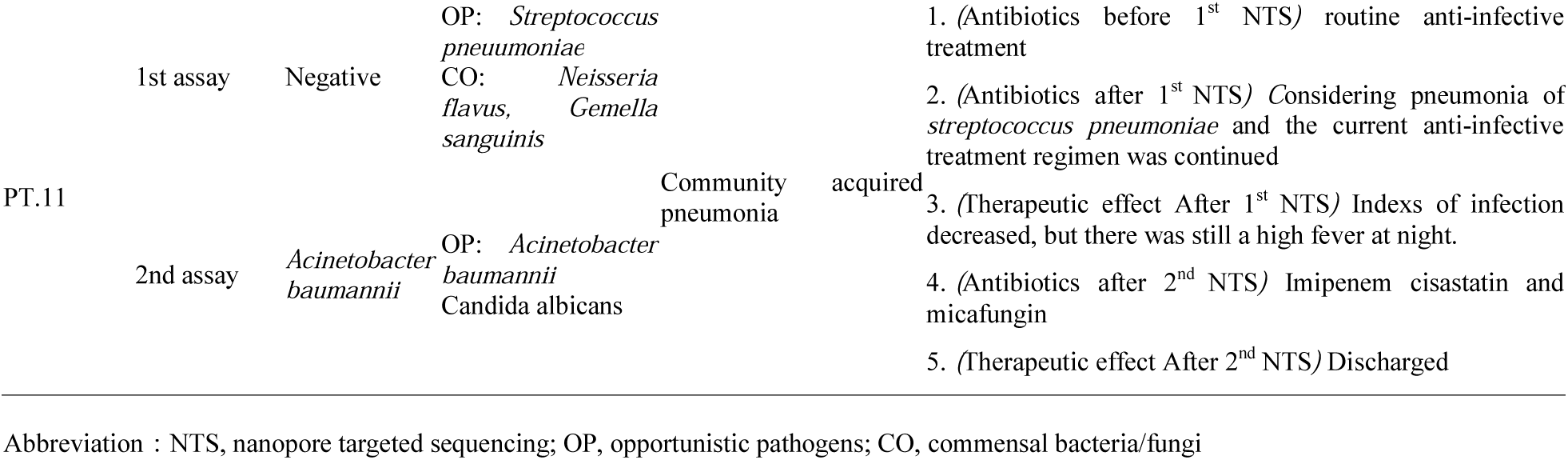
Continuous NTS assay during the treatment process.

## Discussion

We developed and examined the performance and clinical impact of targeted full-length amplicon sequencing in clinical practice, in a setting where most patients have negative culture results *(8)*. NTS increased the sensitivity of pathogen detection for patients with diagnosed or suspected infection and detected more causative pathogens in patients with a risk of infection. These findings are consistent with those of previous sequencing-based studies *(6, 8, 30, 31)*. Compared to culture methods, the detection rate by NTS was higher for nearly all specimen types, particularly in the cerebrospinal fluid (CSF) and aqueous humor. Moreover, slow-growing, uncultivable pathogens were detected in a single NTS test, particularly fungi and MTB *(32)*. For infections induced by culturable bacteria, NTS was less impacted by pre-antibiotic exposure and more effectively detected coinfections. Potential pathogens were detected in specimens with no clone growth in culture. Continuous NTS helped clinicians determine effective treatments, suspend, or de-escalate the use of targeted drugs, and monitor the risk of secondary or nosocomial infections.

By NTS, a turnaround time of <24 h can be achieved for all specimens. Given its high NPV using culture results for reference, a negative result or non-bacterial etiology can lead to de-escalation or even the discontinuation of antimicrobial therapy *(33)*. Moreover, a 6-h diagnostic pre-report can be issued if diagnostic thresholds are met. We encountered emergency cases for which this pre-report substantially improved the diagnosis and care. We identified *Nocardia farcinica*, which is rare in China, in a patient with an emergency intracranial infection, and extremely fast-growing pathogens in the tissue of a patient with Paget’s disease. In addition, NTS detection of fungi and rare coinfections of fungi with symbiotic bacteria, often neglected by routine clinical diagnosis, is possible.

Our results clearly support the combination of targeted amplification and nanopore sequencing; however, amplification may alter the flora structure, affecting the detection of pathogens with ultralow abundance. A total of 80% of NTS false-negative samples were derived from the respiratory tract, suggesting that the extremely high proportion of commensal colonizers by improper sample procurement impacted the sensitivity of NTS. NTS amplification requires adjustment depending on the specimen type. Importantly, many potentially pathogenic bacteria, such as *Pseudomonas* and *Streptococcus*, are ubiquitous *(27)*, and their abundance must be contextualized by cohort-specific norms.

This study had several limitations. First, it was conducted at a single central hospital, and the number of specimens varied substantially among departments. Further studies at hospitals with different patient populations and medical practices are needed. Second, unmeasured factors may have influenced decision-making. In addition, we cannot measure the impact of NTS on patient outcomes beyond antibiotic selection. Finally, given the retrospective nature of the study, all available tests were not performed on all samples, and we did not systematically compare NTS results to those of other molecular diagnostic methods. Future studies should focus on the impact of NTS on patient-centered outcomes (i.e., length of stay and mortality), antibiotic stewardship, and cost compared to a wider range of methods.

In conclusion, the NTS approach developed in this study enabled simultaneous detection and same-day reporting of a myriad of bacteria and fungi in specimens with practical operability in clinical settings. This approach fills the gap between highly targeted PCR-based diagnostic methods and resource-intensive sequencing-based metagenomics methods. The equipment required for NTS testing is small and portable; therefore, NTS can be directly performed in various hospitals or clinics, without sending specimens to large sequencing institutions or third parties detection institutions. NTS is recommended for monitoring severe patients during treatment because of its accuracy, comprehensiveness, and rapidity.

## Data Availability

All data, models, and code generated or used during the study appear in the submitted article.

## Abbreviations

NTS: nanopore targeted sequencing
ITS1/2: fungal internal transcribed spacers 1 and 2
ATCC: the American Type Culture Collection
MTB: *Mycobacterium tuberculosis*
BALF: bronchoalveolar lavage fluid
CFU: colony-forming units
RPM: aligned reads of per million sequence reads
CSF: cerebrospinal fluid
NTM: *Nontuberculous mycobacteria*
PPV: positive predicted value
NPV: negative predicted values
OP: opportunistic pathogens
CO: commensal bacteria/fungi

